# Deep Sit: a system for studying and exam preparation

**DOI:** 10.1101/2025.10.29.25338960

**Authors:** Ahmed Bassiouni, Chi-Kee Leslie Shaw

**Author notes:** **Corresponding author:** Ahmed Bassiouni, Department of Otolaryngology, Head and Neck Surgery, Northern Adelaide Local Health Network, Modbury Hospital, Smart Rd, Modbury, SA 5092, Australia. **Disclosures:** The authors have no financial interests to disclose. **Data Availability statement:** The code for the Deep Sit Obsidian plugin could be found at https://github.com/bassio/obsidian-deepsit.

## Abstract

Studying for medical exams is a great undertaking traditionally considered a “rite of passage”, as the depth and breadth of knowledge required increases progressively from undergraduate-, to postgraduate-, to fellowship-level exams. A common theme of these exams is the necessity of learning and committing to memory a large body of work, corroborating multiple disparate primary sources. The article presents new software, written to assist exam candidates with the process of knowledge acquisition and exam preparation. The software is written as an open-source plugin of the popular free note-taking software Obsidian and provides tight integration with the open-source reference management software Zotero. We then present a study system termed “Deep Sit” which incorporates a simple heading-based organization of medical knowledge: (Definition; Epidemiology; Etiology; Pathophysiology; Symptoms & Signs; Investigations; Treatment). We demonstrate how the Deep Sit study system is significantly facilitated through a walkthrough of these software components.

## Introduction

Preparing and studying for medical exams is a great undertaking. These exams are a source of significant stress for candidates, but have been traditionally viewed as a “rite of passage” to mark a standard of competency and peer recognition, allowing further career progression. Whichever format these examinations come in, they all share a common objective in the form of the learning experience. Learning styles naturally differ from candidate to candidate and are partially dictated by the format and requirements of the exam. A common theme however remains, and that is the necessity of **learning and committing to memory a large body of work, corroborating multiple disparate primary sources**. The size of the curriculum required to study correlates with the level of exam, and thus increases significantly from the pregraduate, to post-graduate, to fellowship (board-level) exams.

There are multiple publications in the medical literature offering high-level advice on exam preparation(1–3). There are also numerous books that offer summarized curricula or synthesized “study notes” for the study of certain exams, each of them usually focusing on a particular exam (these are typical for licensing exams, such as the USMLE exams). There are however few articles that discuss the low-level practical aspects of *how to study the required material (i.e knowledge acquisition)*, taking into account the depth and breadth of knowledge required. This is particularly relevant for exams at the fellowship or doctorate level, in which a deep knowledge and synthesis of the underlying medical literature and evidence is absolutely required. We believe this is (at least in part) due to the fact that studying and learning styles are highly individualistic and could vary tremendously from person to person.

Digital technology has the potential to greatly assist in the process of studying. The reported educational benefits of digital technology are mostly concerned with the “logistics” and “time-saving” aspects of study.(4) On the other hand, digital technology could act as a negative source of distraction, reducing the efficiency of the studying process.(5)

In this paper, we present new software, which had been originally written by the first author (A.B.), specifically to assist with his fellowship examination preparation. The software is written as a plugin of the popular free note-taking software “Obsidian”(6), and provides tight integration with “Zotero” which is one of the most popular open-source reference management software packages.(7) The article then presents an introduction to a study system termed “Deep Sit”, which is greatly facilitated by these software components. The article illustrates this within a medical setting, but the principles could be transferred to other academic disciplines.

## Requirements

Requirements for a study system for an important exam are highly personal. These will typically vary with the learning style of the individual and the requirements of the specific examination to be sat. Presented here are a personal account of study requirements that led to development of Deep Sit.

### a) Effective note-taking

Note-taking offers multiple well-established, evidence-based advantages.(8) The first advantage is having notes available long-term for efficient later review; this is known as the *external storage effect* of note-taking. The second purported benefit is that the process of taking notes itself engages the individual in a deeper level of information processing and comprehension, which is known as the *encoding effect* of note-taking.(8)

There is suggestion in the literature that utilizing one’s own written notes may carry additional benefit when compared to, for example, using borrowed notes from others.(9) This could be related to the “depth of processing”(10) employed during the process of writing and formatting one’s own notes, in their own preferred learning and recall style, and is related to the semantic encoding effect. Or alternatively this could be explained in terms of time spent processing and revising.(9) Despite this, there is some evidence that lecture-provided notes adds a significant effect size to the benefits of note-taking and note-reviewing.(11) This suggests that note-reviewing has the greater effect size when compared to note-taking.(11) Obviously, writing one’s own notes (from scratch) entails additional time investment and effort. Ultimately, this choice will depend on the individual’s learning style and it is recommended for each exam candidate to utilize the note-taking strategy that would maximize their knowledge retention and recall.

### b) Reducing overhead from non-essential tasks including formatting

Digital note-taking offers additional advantages in regards to note-taking efficiency compared to traditional handwritten notes, and is now considered widespread practice in multiple settings.(12) This calls for the use of efficient note-taking software that does not detract from learning and deeper processing during. Formatting, organizing and polishing notes are some of these important tasks that nevertheless could detract from learning during note-taking.

Obsidian is one of the popular free markdown-based text editors that are commonly used for note-taking. Obsidian naturally takes advantage of the readability of markdown format, but its main advantage in ease of note-taking is its WYSIWYG (What you see is what you get) principle. Markdown WYSIWYG means that formatting is done automatically as typing is performed, allowing well-organized and good-looking notes by default while minimizing distraction and time investment.

### c) Ability to organize notes in both hierarchical and non-hierarchical fashion

Many medical disciplines could be easily organized in the form of body systems or subspecialties. The hierarchical folder structure allows easy and quick localization to notes, constituting a “map” of sorts. Another additional technique for organization is the utilization of “tags”. Tags allow labelling grouping of multiple notes (sometimes in separate folders or hierarchies) under a certain tag in a non-hierarchical fashion. The preference to use folders versus tags for organization may differ based on the subject matter and the preferences of each individual. The combination of folders and tags arguably offers the most versatility.

### d) Accessibility to notes from different devices and the “local-first” principle

Keeping a copy of the notes locally on one’s own device was deemed a significant advantage, rather than only keeping one copy in the cloud. This is the “local-first” software principle.(13) The main advantage of this is that one does not need an internet connection active all the time while studying or editing their notes. As such, multiple copies of your notes should exist, one on each device. This redundancy is also important as a backup mechanism. The software used (or other software synchronization plugins or solutions) should allow syncing the study material across all devices to keep any changes up to date. This is crucial for maximizing study time, particularly while working full-time. At home, one could be using your desktop or laptop for study, but at the hospital/work the laptop may not be readily available all the time and thus the need to use a work based PC for study and research. Having rapid access to notes on one’s mobile phone is also an additional advantage.

### e) Effective reference management with highlights and annotations

We use Zotero, a free and open-source reference management software created at the Roy Rosenzweig Center for History and New Media at George Mason University.(7). It is one of the most capable and advanced software packages for reference management. There are other capable reference managers including Mendeley and EndNote, but they are either not open-source (the former) or not free (the latter). Zotero is rare in that it allows for the installation of custom user-programmed plugins to enhance its functionality. This is essential to allowing integrations with other software (including note-taking software such as Obsidian) to be developed.

Zotero allows storage and organization of references in its library into folders and subfolders (termed “Zotero Collections”). This allows the user to customize the folder hierarchy of where each reference is stored, allowing for easier retrieval. Any reference item could also be added to multiple Collections. There is also a tag system to allow non-hierarchical methods of grouping items.

In addition, Zotero has its own integrated pdf and epub reader, thus allowing direct review of the reference’s full text when available without the need for an external viewer. The newer versions of Zotero allows taking annotations (i.e. highlighting important information) in the form of specific areas of text or images within the reference’s pdf file and storing these within Zotero’s database and accordingly synchronized across devices.

### f) Easy review of primary sources

Easy access to primary sources during note writing was deemed essential. This depth and level of knowledge of the literature and the underlying evidence behind medical practice is not usually required at the Undergraduate level, but is a prerequisite at the Fellowship or Doctorate level.

Relevant highlights and annotations should be easily viewable during study of a certain topic; “Going back” to consult the original sources during study should be facilitated. Since this requirement is not attainable within Obsidian alone (since it is not a reference management software), a new plugin or integration was required.

## Deep Sit: a study system for medical knowledge bases

We introduce a study system termed “Deep Sit” that incorporates a simple heading-based organization of medical references and knowledge. We derived this name from an acronym “DEEPSIT”, based on the list of common headings that allows one to **summarize the core body of knowledge related to a certain disease or condition**. (Table 1) These headings are probably familiar to many undergraduate medical students, as they are commonly used for teaching in medical school.

**Table 1:**
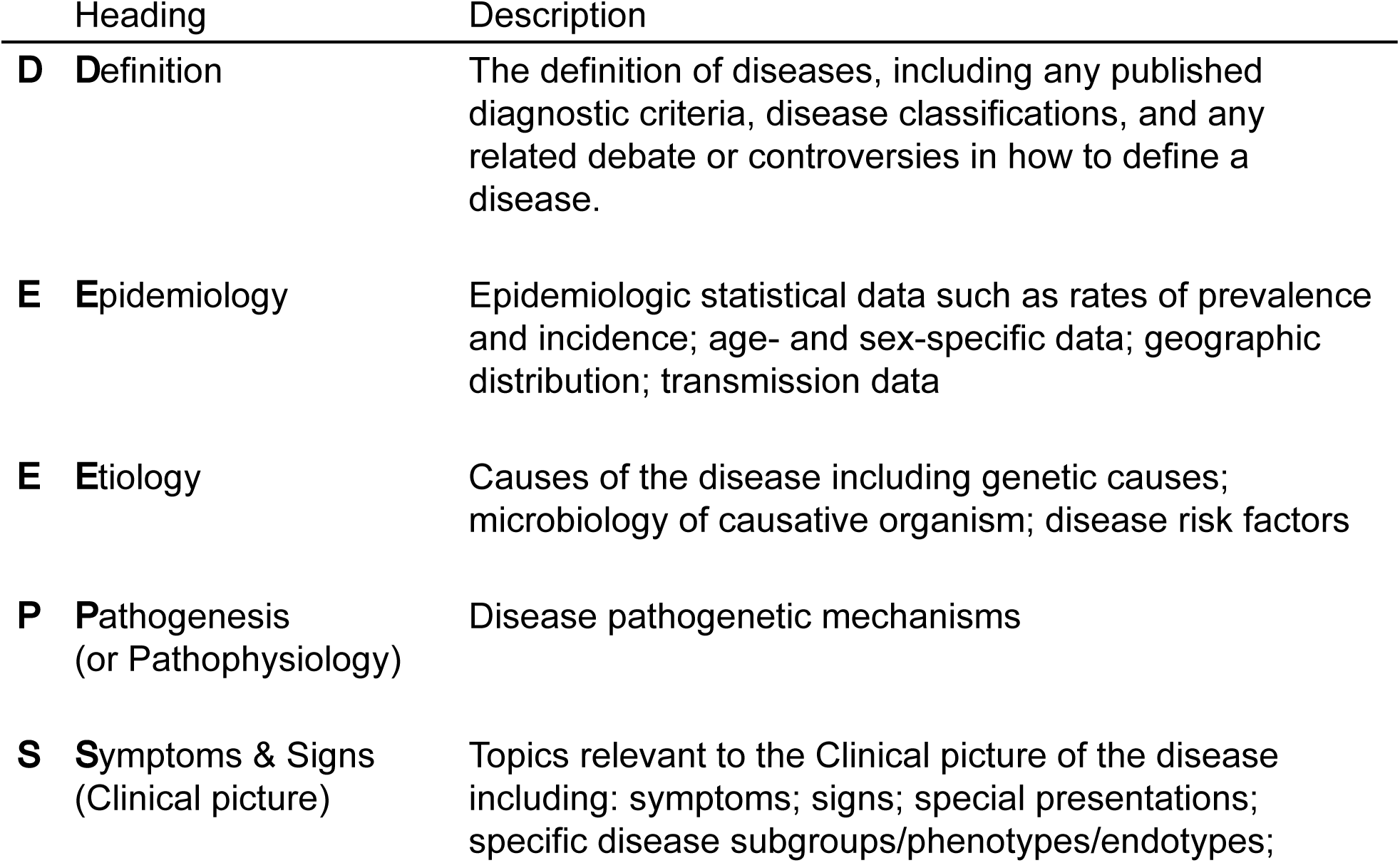
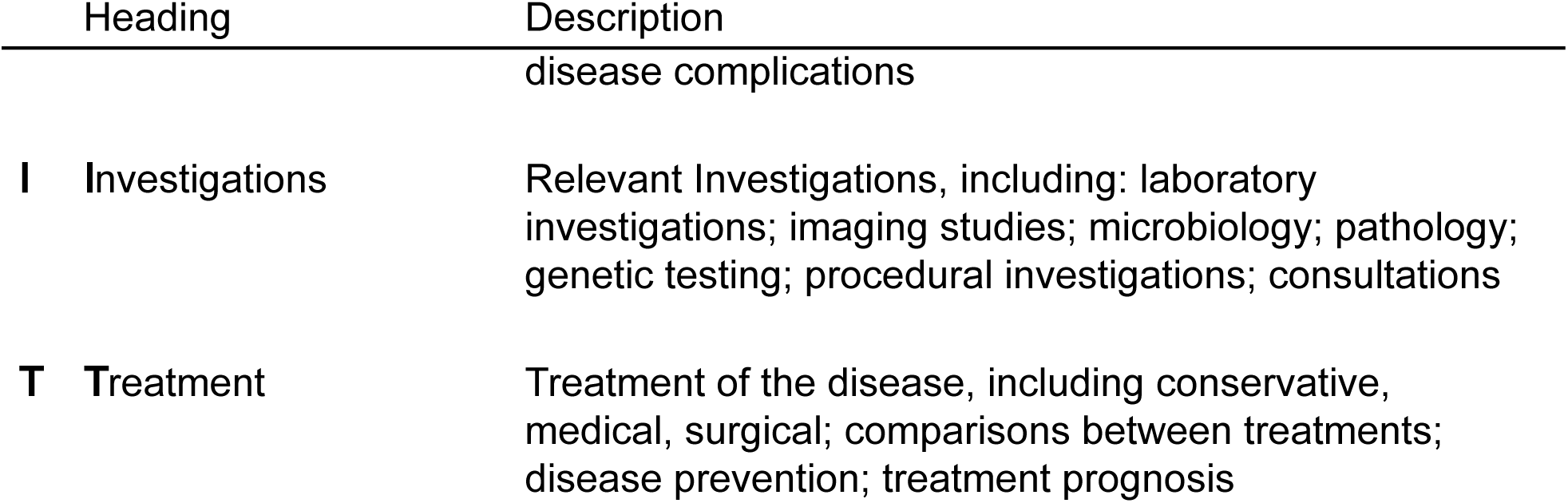
*The Deep Sit system is* used for a simple heading-based organization of medical knowledge.

We also find the acronym memorable, as it acts as its own mnemonic, reminding the students of their main task! Exam candidates repeatedly need to have a “*deep sit*” and study, sometimes gruellingly for hours on end.

## Description of our Obsidian plugin

The first author (A.B.) wrote an Obsidian plugin termed Deep Sit (https://github.com/bassio/obsidian-deepsit), named after the DEEPSIT acronym (as learned from the senior author L.S.). The plugin is written in Typescript, similar to Obsidian. Development of the plugin commenced in December 2023, initially as a replacement for in-text bibliography citation functionality from Zotero. It however later expanded in scope to include more functionality integrating between Obsidian and Zotero.

The plugin at the time of writing has the following features:

### 1) Integration with Zotero

Zotero has to be running for the plugin to function, along having the Better Bibtex plugin by Emiliano Heyns installed and activated.(14) The Better Bibtex plugin is an important plugin that enhances the management and export of references from within Zotero and therefore makes it easier to manage bibliographic data for text-based workflows.(14) The Better Bibtex plugin allows the Deep Sit Obsidian plugin to communicate with Zotero and extract the required bibliography and annotation information.

### 2) Bibliography citation

Taking advantage of the Zotero integration above, provided by the Better BibTex plugin, the Deep Sit plugin allows searching a Zotero Collection (as indicated in the note’s YAML metadata) for bibliography entries and citing these in the text (i.e. “cite-as-you-write” with a suggestion dropdown list). Citations are enclosed within one pair of square brackets with semicolons separating single items (+/- spaces). Each reference is cited using its “Citation key” as set up by Zotero / Better Bibtex, preceded by the ‘@’ sign. This citation syntax is in accordance with Pandoc’s citation syntax.(15) An example is provided below:

~~~
these patients do better with Bone Conduction Devices than with conventional hearing aids.[@Mylanus1998; @DeWolf2011]
~~~

### 3) Reference list features

The plugin allows keeping track and visualizing the references contained in the related Zotero Collection alongside the note’s markdown in Obsidian’s right-handed sidebar. There are two modes for this list. The default mode is termed “References mode” which presents an ordered list of only the reference items cited within the text. A second mode is termed “Bibliography mode”, and this alternatively allows to list all the References contained within the accompanying Zotero Collection, not limited to the ones cited within the text. Whichever mode is chosen, Zotero references that have a pdf attachment available (i.e. the full-text of the article) are highlighted as a link, which when clicked, opens the pdf directly in Zotero’s reader for direct viewing.

### 4) Highlights and annotation features

The Deep Sit plugin highlights references in the sidebar that contain highlights and annotations and provides an efficient “Multi-annotation” viewing feature, that lists all text and image highlights and annotations for all pdf files within a Zotero collection. This allows review of previously studied sources, and rapid viewing of underlined or highlighted texts alongside one’s study notes, without the need to keep switching between different applications.

## A walk-through of the Deep Sit workflow

An example walk-through for a Deep Sit study session that uses the above software components is provided below in the context of the field of Otolaryngology, Head and Neck Surgery (also known as Ear, Nose & Throat, or “ENT” for short).

The medical condition “Meniere’s disease” is chosen as the topic for illustration. Menière’s disease is a disease of the inner ear associated with cochleo-vestibular changes of endolymphatic hydrops (increase in pressure of the endolymph). Symptomatically, it is characterized by episodes of spontaneous vertigo, usually associated with unilateral fluctuating sensorineural hearing loss, tinnitus and aural fullness.

### Reference management: Zotero

Pubmed, Google, or Google Scholar (and other online reference databases) are great online avenues for initial search for papers on a certain topic. AI assistants such as Anthropic’s Claude or OpenAi’s ChatGPT will also be very relevant for this kind of search in the future, although currently the risk of “AI hallucinations” limits their use in academic settings, particularly when asking for primary references. General medical journals such as NEJM, JAMA or the Lancet provide high level narrative reviews of disease, otherwise specialty journals are very valuable in that aspect. Add the relevant papers into Zotero’s library; either manually through the reference’s DOI, or through the various Zotero browser connectors. Zotero is a well established software package; Numerous Zotero tutorials exist online and a full rundown of features or a “How-to” is out of the scope of this article.

Make sure to place the reference in an appropriately-named collection (in this example, “Meniere’s disease”). From a folder organizational point of view, this collection could be placed as a subcollection under the Collections “Otology -> Inner ear disorders -> Meniere’s disease”. Full texts of these references are sought and downloaded, and could be added to Zotero’s storage. Using Zotero’s built-in pdf reader, read and study the primary sources in keeping with your learning objectives, making sure to highlight important text or images and add annotations for later review. Search interesting references, fetch their full text, and add these to your reference collection as you see fit.

As the depth of research and knowledge increases, references will continue to accumulate under the “Meniere’s disease” collection/folder. There will then come a need to add “sub-collections” inside the top level collection, and move/copy the relevant references under these subheadings for better organization. For consistency, we recommend that the scheme used for naming the sub-collections could follow the Deep Sit classification (Table 1) at the top-level folder. (see Figure 1 for an example)

**Figure 1:**
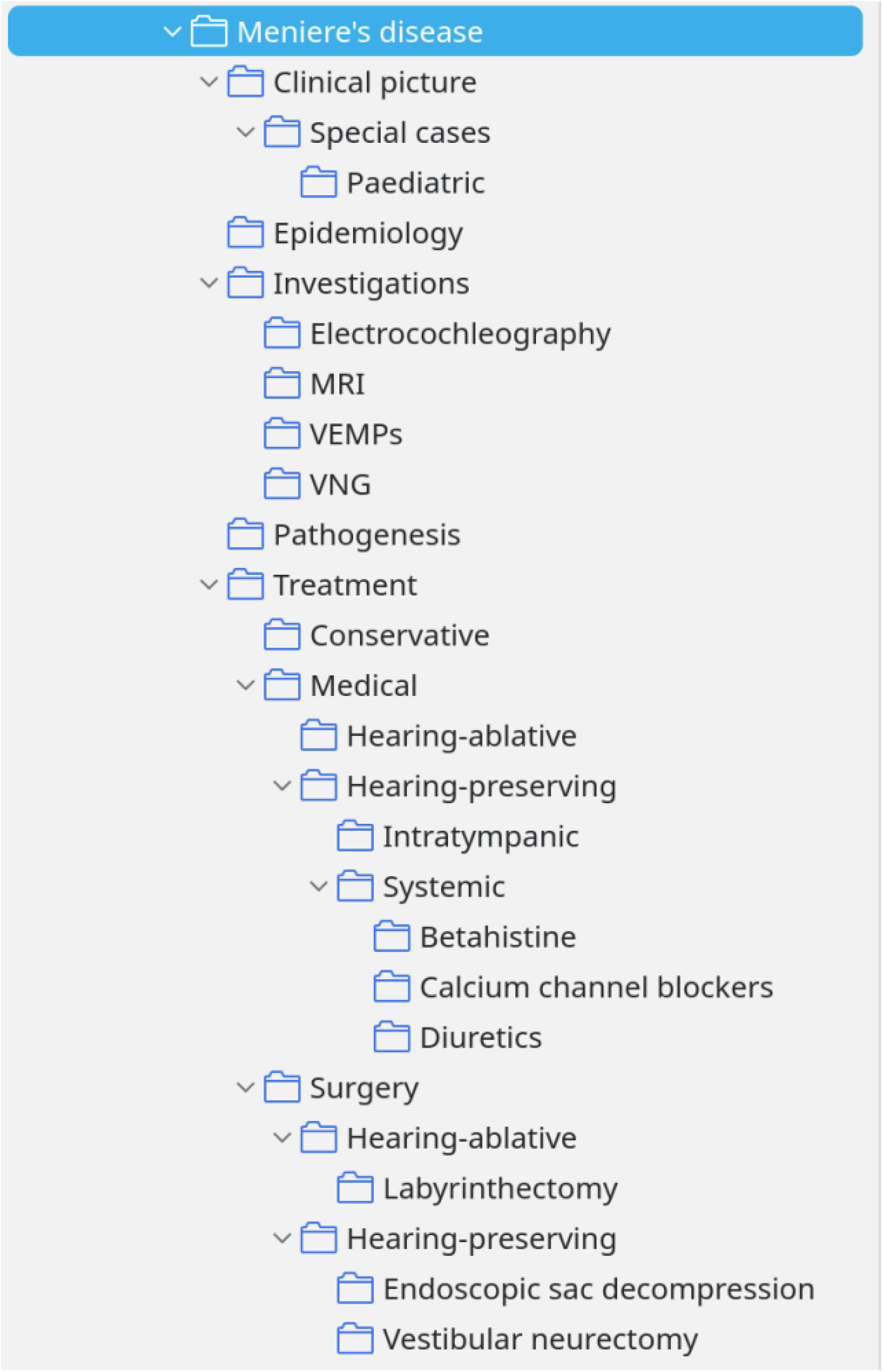
An example of a DEEPSIT scheme applied to “Meniere’s disease” Collection Zotero refernce manager.

As per the example noted in Figure 1, the Deep Sit classification will be very useful for the user once the reference library grows beyond a certain size.

### Note-taking: Obsidian

Download and install Obsidian according to their help manual (https://help.obsidian.md/install). Obsidian organizes its knowledge bases in the form of “Vaults” which is essentially a folder on the local file. Obsidian is able to work on one Vault in any one open Obsidian window at a time. Each vault/folder could contain folders and subfolders (in addition to the actual “notes”). Some users online report preference for a “flat-folder hierarchy” and use tags for organization. While this is user-dependent, we do not advocate for this system in the setting of a fellowship exam, as we believe that personal hierarchical organization is very important for rapid folder navigation.

Each note in the vault is represented by a markdown file with the “.md” extension. MD files are ultimately text files and are viewable as simple text, without formatting, by any other text viewer, including notepad. Place the notes under the appropriate folder hierarchy, according to the curriculum.

For example, a subspeciality-based top-level folder structure for ENT may include the following: - General ENT - Paediatric ENT - Otology - Rhinology - Laryngology - Head & Neck - Facial plastics - Sleep Medicine - Audiology - Anatomy - Pharmacology - Physiology & Pathology

Folder structure is fluid and can be changed anytime, therefore one does not need to worry too much about the exact initial folder structure, as this is expected to continue to change and expand with time as the knowledge base grows.

After the preliminary folder structure is created, commence by creating a new Note in Obsidian under the intended folder (right-click -> New note).

The template functionality of Obsidian could be used to quickly insert ready-made repeatable patterns of text in a note. (see Figure 2 for an example template that incorporates the DEEPSIT headings)

**Figure 2:**
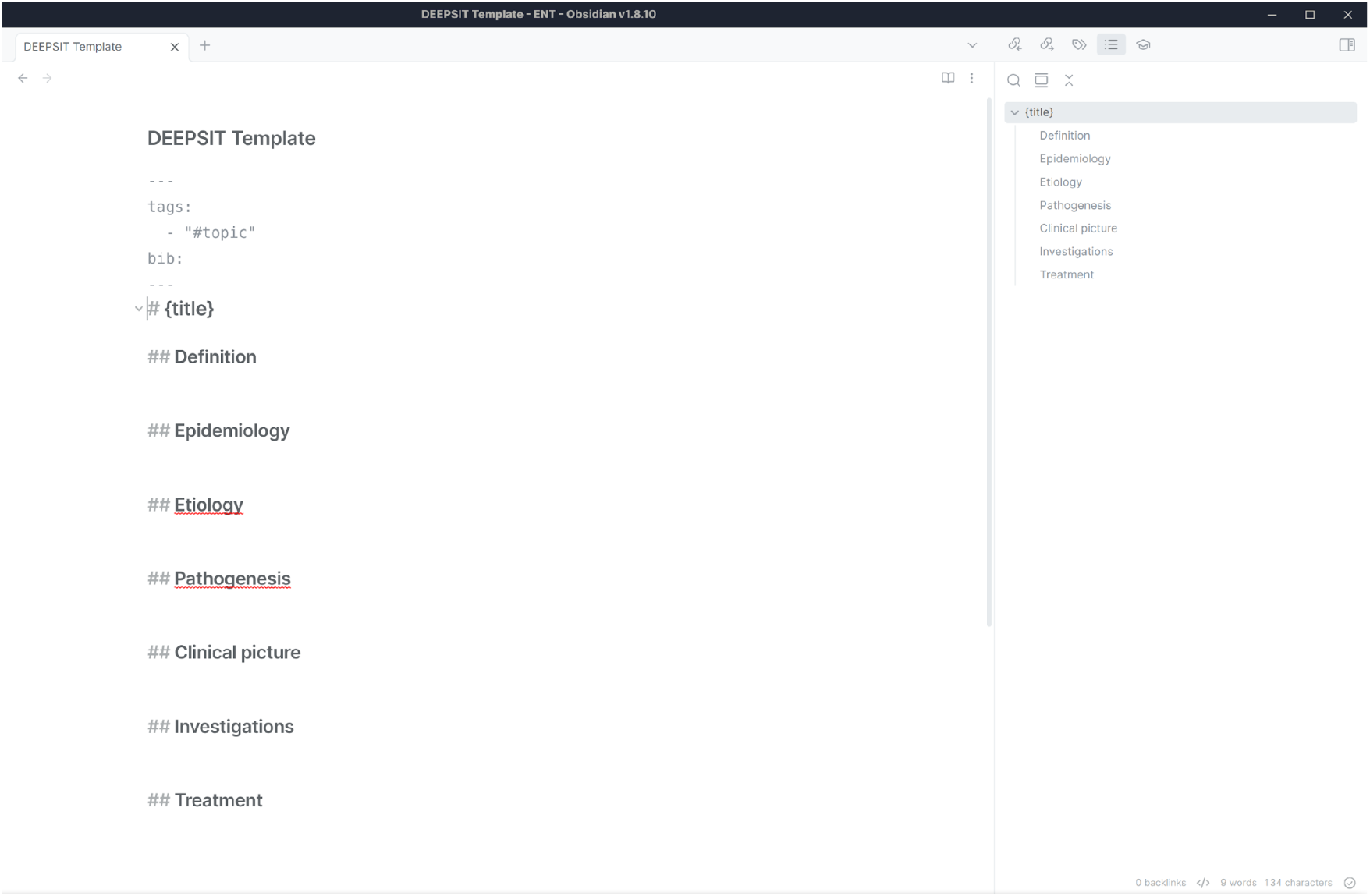
Example of a DEEPSIT template in Obsidian.

How does one write and format their notes in Obsidian? Note-taking in Obsidian differs from the sophisticated capabilities of word processors such as Microsoft Word. Obsidian uses a simple and widely used markup format, called “Markdown”, which we will briefly introduce below.

### Introduction to the Markdown format

Markdown format is a widely used simple text writing format. It was first proposed by John Gruber in 2004.(16) Since then, multiple slightly different formats or “flavours” have been described, but they all follow the same principles as laid down by Gruber. Standardized markdown efforts include the CommonMark format(17), the Pandoc-flavoured markdown(18), or Github-flavoured markdown.

The principle philosophy underlying markdown is easy **readability** by a human reader, as it employs self-describing formatting indicators. This allows the files to be viewed using simple text editors such as notepad, without requiring a word processor. Very low additional time investment is thus required for formatting notes and keeping them in a presentable format. This is particularly advantageous in exam preparation settings.

Markdown is widely known and utilized in information technology (IT) circles, but may not be familiar to those from a medical or biomedical background. The most frequent features of markdown will therefore be presented to the readers below, however full rundown of its specification is not within the goals of this article.

#### Emphasis and simple formatting

Italic characters (also known as ‘emphasis’) are marked by enclosing the italicised text between a pair of single asterisks (*) or underscores (_). Bold characters (or ‘strong emphasis’) are marked by enclosing the highlighted text between double asterisks (**). Text that is intended to be both italicised and emphasized is typed between triple asterisks (***). These emphasis characters could be used in the middle of a word too.

~~~
This line contains *italic* text and *part*ially italicised words.
This line contains **bold** text.
This is both ***bold and italic***.
~~~

#### Headings

Headings (and sub-headings) are commonly indicated via hash character(s) at the beginning of the line. The number of hash characters at the beginning of the line indicates the level of the “sub”heading (up to six levels allowed).

~~~
# First-level header
## Second-level header
### Third-level header
~~~

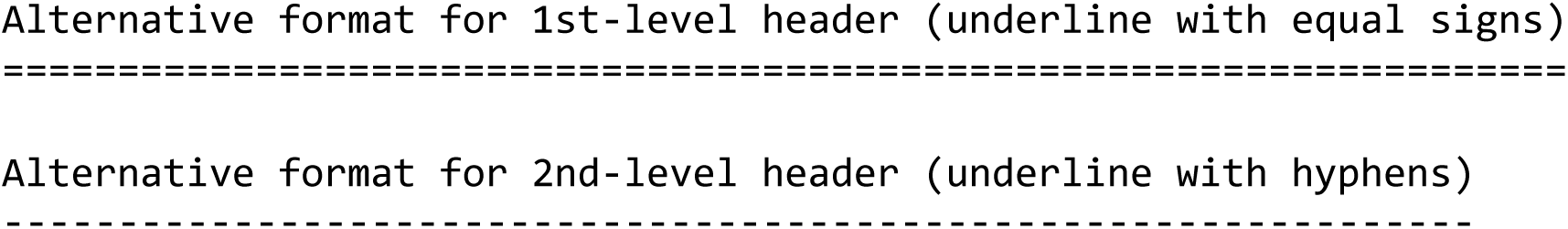

#### Paragraphs

A paragraph in markdown is composed of one or more consecutive lines of text, separated by one or more blank lines (i.e. lines that contain nothing, with the exception of spaces). This has the implication that a paragraph could be written as a separate list of sentences one after another on separate lines, without indicating a start of a new paragraph.

Intentionally inserting a “line break” (i.e. an instruction to force a new line at the end of a line of text) requires inserting two spaces at the end of the line after the text ends.

Paragraphs could also be appended underneath list or blockquote items. (See below)

#### Lists

Perhaps the most frequently used features are lists. Lists could be either numbered (i.e. ordered) or un-numbered.

Un-numbered lists are marked at the beginning of a new line using either a hyphen (-), an asterisk (*), or a plus sign (+).

An example of a list:

~~~
– List item x
– List item y
– List item z
~~~

Numbered lists commence with a number followed by a period “1.” or a closing bracket “1)”

A numbered list example:

~~~
1. List item number one
2. List item number two
3. List item number three
~~~

Lists could also be indented, and (as mentioned above) could contain multiple lines / child paragraphs. If list items are separated by blank lines, the markdown processor will wrap the list item’s text content in a paragraph tag.

~~~
– item
 –indented subitem
– second item
 –indented subitem with multiple lines. second line.
–third item
~~~

#### Links

A hyperlink is indicated between square brackets for its text, followed by curved brackets containing the link’s URL.

~~~
[This is a link](http://example.com) to a website.
~~~

Markdown also supports a different format called reference-style links, where the link’s address and title are indicated separately (for example: at the end of the text, similar to a reference list).

Obsidian also natively supports another useful link format. This link format is not included in the official markdown specification, but it is a widely-used linking format first popularized by Wikipedia. This format is called “wiki-style links” and are commonly used for internal linking. This is a very valuable short-hand for linking one note to another in Obsidian.

~~~
# Parapharyngeal space tumours
## Definition
Tumours occurring in the [[Parapharyngeal space]].
~~~

In the example above, a note called “Parapharyngeal space tumours” provides a wiki style link to another note, the “Parapharyngeal space” where for example the anatomy of the parapharyngeal space could be quickly reviewed during a study session.

#### Images

The syntax for inserting images is identical to that of links (see above) but preceded only by an exclamation mark “!”. Here the image’s description (or the “Alt text” of the image) is enclosed between the square brackets, while similar to links the path to the image file or URL is enclosed between the curved brackets.

Example image:

~~~
![Alt text](/path/to/img.jpg)
~~~

#### Horizontal rules

A horizontal line or ruler could be inserted by typing three or more hyphens (—), asterisks (***), or underscores (__) on a line by themselves.

#### Full markdown specification

The full markdown specifications could be reviewed online at multiple sources. (17–19)

### Integrating note-taking with primary sources: the Deep Sit plugin

Install the Deep Sit plugin manually into your Obsidian vault by following instructions from the github repository. Enable the Deep Sit plugin through the Community plugins tab through the Obsidian settings and set up the plugin’s settings.

Once the plugin is enabled, a new icon should appear in the right-handed sidebar which visualizes the Deep Sit References View when clicked. (Figure 3)

**Figure 3:**
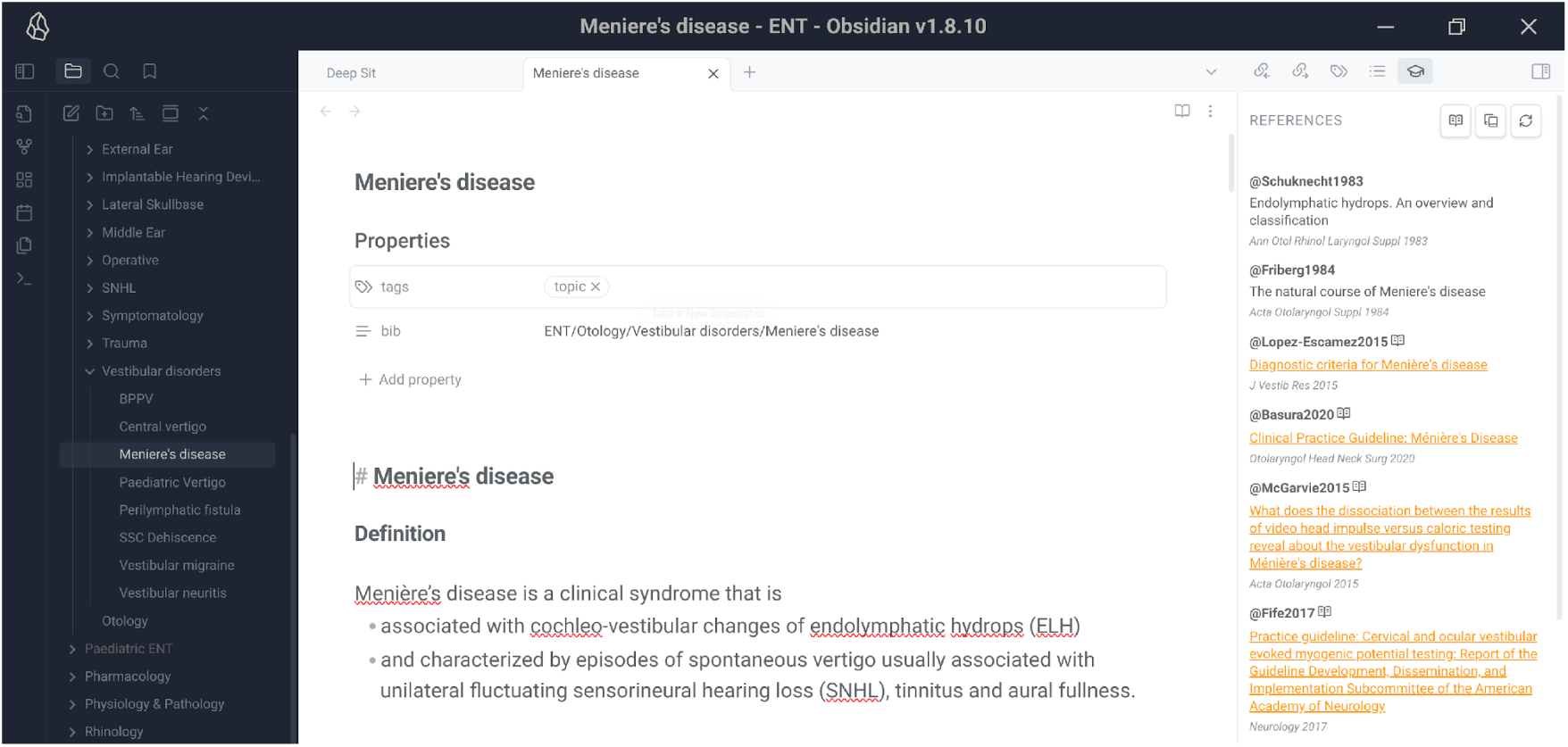
Screenshot demonstrating the Deep Sit plugin’s sidebar view of a reference list for a markdown file.

The next step is to connect the markdown note to a specific Zotero collection. This is indicated via the file’s YAML metadata present at the top of the file (also called “Add file property” in Obsidian). Add a new property with the title “*bib*” and a value that corresponds to the path to the appropriate Zotero collection/folder, for example: *My Library/Otology/Vestibular disorders/Meniere’s disease*. This indicates to the plugin that the current note being edited should consult the “Meniere’s disease” collection within the Zotero library called “My Library” and located under “Otology -> Vestibular disorders”. Once this is entered, pressing the Refresh button in the Deep Sit right-handed pane should perform a refresh and refetch the bibliographic entries for that Collection from Zotero. This also fetches any pdf highlights and annotations for the bibliographic entries, which are viewable via clicking the Annotations button in the plugin pane.

The integration allows the list of references to be visible in the right-sided sidebar. (Figure 3) This connectivity to Zotero requires the Better Bibtex Zotero plugin(14) to be installed and active, and Zotero to be open and running in the background.

Once confirmed that the plugin successfully fetched the relevant bibliography from Zotero, inline citations could also be queried and entered into the text while typing. To add citations to the markdown note, use the Pandoc citation format “[@Citationkey]”.(15) A pop-up suggestion dropdown list will appear to allow selecting the appropriate reference. (Figure 4)

**Figure 4:**
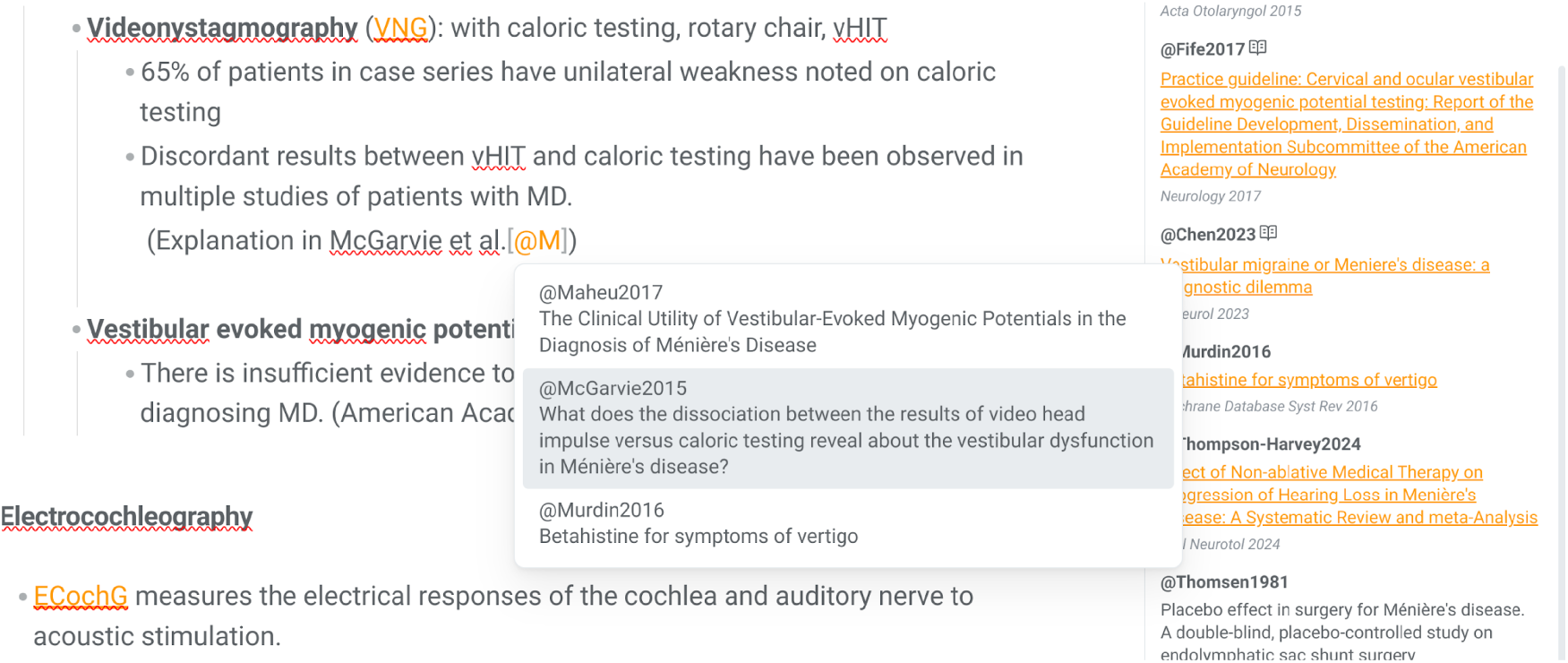
Screenshot demonstrating the “cite-as-you-write” functionality via querying Zotero.

Finally, during later note-review for purpose of study, the plugin allows an immediately-available reference list along with previously highlighted texts and figures from the original texts on the right-hand side of the screen. (Figure 4) If further consulting of the original pdfs is required, then it is accessible via single-click that jumps to the original highlights in the pdf.

**Figure 5:**
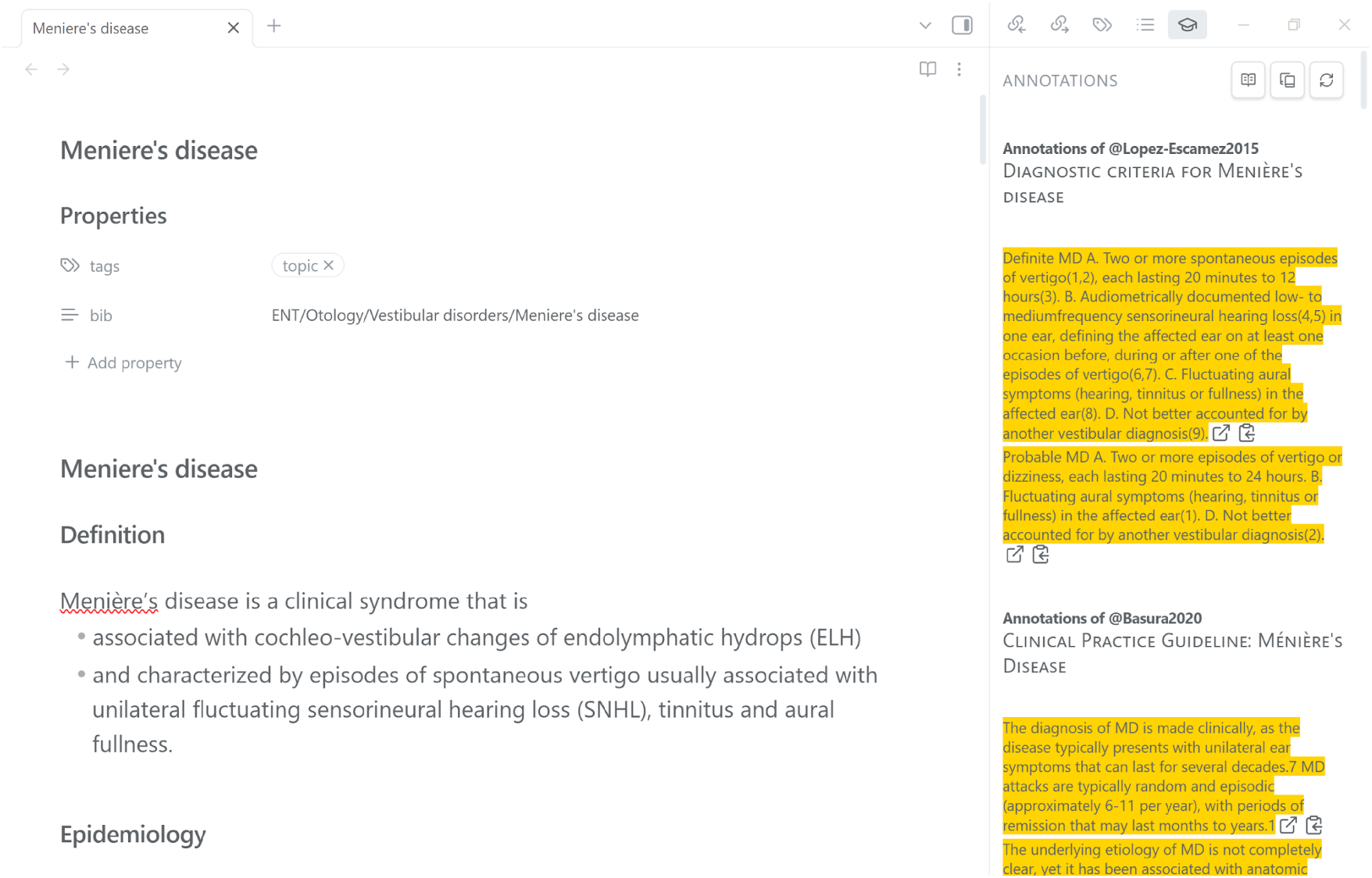
User highlights or “annotations” are easily accessible via the sidebar.

## Discussion

Sitting examinations is a rite of passage in many professions and disciplines. In the medical setting, multiple articles have tackled this topic, providing general advice and specific tips for exam preparation. Here we provide a personal account that assists with study through a digital workflow that utilizes a new plugin that integrates note-taking and reference-management software.

We presented a practical walkthrough through a study system that involves maintaining a library of primary sources, as well as maintaining a personal knowledge base in the form of typed notes, using existing free software; the *Deep Sit* system is an easy-to-remember and familiar scaffold to notate medical knowledge and to organize reference material. The new obsidian-deepsit plugin was then authored to integrate the note-taking software (Obsidian) and reference-manager (Zotero) to provide a focused and effective study and research tool.

Similar software solutions to our plugin currently exist, but none to our knowledge that provide this level of integration on a per-file basis. First, for Obsidian, multiple Zotero integration plugins(20) with similar goals to our plugin already exist, but they utilize different workflows and some have more advanced functionality. The closest functionality to our plugin we found in the excellent software package Zettlr(21), an academic open source markdown editor which provides a similar WYSIWYG editing feature as Obsidian, with in-built citations and a sidebar view of reference lists. However, Zettlr does not appear to provide support for displaying stored highlights and annotations at this stage.

There are several limitations to the study approach presented in this article. First, is that it is highly personal and therefore may not suit all individual study styles. Second, it is a purely digital workflow in that there is currently no way to incorporate information locked in non-digitized textbooks and hand-written notes (other than to transcribe them manually into the digital knowledge base). The literature is currently ambiguous about the superiority of digital over traditional note-taking. Digital workflows are generally viewed as less practical as they lack the simplicity of pen-and-paper, but it is clear they do offer a level of efficiency when compared to the traditional methods. There may be some solutions developed in the future to address this, particularly if further integrations are developed with tablet products that imitate pen-and-paper using a stylus (for example, the reMarkable series of tablets). Thirdly, the workflow could be considered complex, in that it requires the integration and customization of multiple software components (i.e. too many “moving parts”), and hence it will present a steep learning curve for some users. It also involves the need to learn and be comfortable with the markdown format. Some exam candidates may therefore prefer to use only one software package to address their note-taking needs, one that does not involve learning how to take quick notes in markdown format.

Aside from the limitations, it is also important to mention the advantages of our approach. The main advantage of the digital workflow presented here is that it encourages review of primary sources and incorporates it into one’s study habits. At the fellowship level, a common learning objective is to be aware of the literature or evidence that supports medical practice. This level of data processing and research potentially augments consolidation of memory and recall of medical facts. Source evaluation or “sourcing”(22) has been found to contribute to a type of memory termed “source memory”(23), a process also known as “source monitoring”.(24) Source monitoring adds additional context during studying that improves the process of learning.(25,26) Another advantage of the workflow is that it is basically free to use (except if signing up for extra synchronization subscription services through Zotero or Obsidian) and utilizes mostly open-source components. There is also no contraindication for this study system to be replicated by other developers for other software packages in the future.

## Conclusion

We recommend the Deep Sit system (Definition; Epidemiology; Etiology; Pathophysiology; Symptoms & Signs / Clinical picture; Investigations; Treatment) as a highly-personalized study system for advanced medical exam preparation that prioritizes multi-source literature review, maintaining an organized reference library, and a simple yet well-formatted note-taking system. We introduce software recommendations including a new software plugin. Further research should investigate the acceptability of this system for medical or surgical trainees preparing for their fellowship exams, and whether it leads to objective improvements in learning outcomes.

## Acknowledgements

We would like to thank the authors of all the software cited (including their underlying dependencies) for publishing their work as free and/or open-source software. Particular thanks to Emiliano Heyns whose work on the Better Bibtex plugin allowed the integration between Zotero and Obsidian.

## Notes

**Funding statement:** This work is self-funded. The authors did not receive payment or services from a third party for any aspect of the submitted work.

### Competing Interest Statement

The authors have declared no competing interest.

### Funding Statement

This work is self-funded. The authors did not receive payment or services from a third party for any aspect of the submitted work.

## References

1. Schattner P, Kidd M. How to prepare for the FRACGP exam. Aust Fam Physician. 1994 Mar;23(3):408, 411–2, 416 passim.

2. Bhuiyan AKMF. Overseas trained doctors. How to prepare for a fellowship exam. Aust Fam Physician. 2004 Sep;33(9):746–7.

3. Stewart SM, Jancevski K, Cocks TP. Surviving the FRACGP and staying sane. Aust Fam Physician. 2004 Sep;33(9):683–5.

4. Henderson M, Selwyn N, Aston R. What works and why? Student perceptions of ‘useful’ digital technology in university teaching and learning. Studies in Higher Education [Internet]. 2017 Aug 3 [cited 2025 Jul 30];42(8):1567–79. Available from: 10.1080/03075079.2015.1007946

5. Patterson MC. A Naturalistic Investigation of Media Multitasking While Studying and the Effects on Exam Performance. Teaching of Psychology [Internet]. 2017 Jan 1 [cited 2025 Jul 23];44(1):51–7. Available from: 10.1177/0098628316677913

6. Obsidian [Internet]. [cited 2025 Jul 15]. Obsidian - Sharpen your thinking. Available from: https://obsidian.md/

7. credits_and_acknowledgments [Zotero Documentation] [Internet]. [cited 2025 Jul 15]. Available from: https://www.zotero.org/support/credits_and_acknowledgments

8. Jansen RS, Lakens D, IJsselsteijn WA. An integrative review of the cognitive costs and benefits of note-taking. Educational Research Review [Internet]. 2017 Nov 1 [cited 2025 Jul 14];22:223–33. Available from: https://www.sciencedirect.com/science/article/pii/S1747938X17300374

9. Carroll K. The Relationship Between Note taking, Revision, and Learning in Tertiary Education: A Review of Literature. All Ireland Journal of Higher Education [Internet]. 2024 Mar 31 [cited 2025 Jul 15];16(1). Available from: https://ojs.aishe.org/index.php/aishe-j/article/view/781

10. Bretzing BH, Kulhavy RW. Notetaking and depth of processing. Contemporary Educational Psychology [Internet]. 1979 Apr 1 [cited 2025 Jul 15];4(2):145–53. Available from: https://www.sciencedirect.com/science/article/pii/0361476X79900699

11. Kobayashi K. Combined Effects of Note-Taking/-Reviewing on Learning and the Enhancement through Interventions: A meta-analytic review. Educational Psychology [Internet]. 2006 Jun 1 [cited 2025 Jul 15];26(3):459–77. Available from: 10.1080/01443410500342070

12. Mueller PA, Oppenheimer DM. Technology and note-taking in the classroom, boardroom, hospital room, and courtroom. Trends in Neuroscience and Education [Internet]. 2016 Sep 1 [cited 2025 Jul 14];5(3):139–45. Available from: https://www.sciencedirect.com/science/article/pii/S2211949316300102

13. Kleppmann M, Wiggins A, Van Hardenberg P, McGranaghan M. Local-first software: you own your data, in spite of the cloud. In: Proceedings of the 2019 ACM SIGPLAN International Symposium on New Ideas, New Paradigms, and Reflections on Programming and Software [Internet]. Athens Greece: ACM; 2019 [cited 2025 Jul 25]. p. 154–78. Available from: https://dl.acm.org/doi/10.1145/3359591.3359737

14. Better BibTeX for Zotero [Internet]. [cited 2025 Jul 15]. Better BibTeX for Zotero. Available from: https://retorque.re/zotero-better-bibtex/index.html

15. MacFarlane J. Citation syntax [Internet]. 2025 [cited 2025 Jul 15]. Available from: https://pandoc.org/demo/example33/8.20-citation-syntax.html#citation-syntax

16. Daring Fireball: Markdown [Internet]. [cited 2025 Jul 12]. Available from: https://daringfireball.net/projects/markdown/

17. CommonMark Spec [Internet]. [cited 2025 Jul 15]. Available from: https://spec.commonmark.org/

18. Pandoc’s Markdown [Internet]. [cited 2025 Jul 15]. Available from: https://pandoc.org/MANUAL.html#pandocs-markdown

19. Daring Fireball: Markdown Syntax Documentation [Internet]. [cited 2025 Jul 15]. Available from: https://daringfireball.net/projects/markdown/syntax

20. Obsidian [Internet]. [cited 2025 Jul 30]. Plugins. Available from: https://obsidian.md/plugins?search=zotero

21. Erz H. Zettlr [Internet]. Zenodo; 2025 [cited 2025 Jul 30]. Available from: https://zenodo.org/doi/10.5281/zenodo.2580173

22. Wineburg SS. Historical problem solving: A study of the cognitive processes used in the evaluation of documentary and pictorial evidence. Journal of Educational Psychology. 1991;83(1):73–87.

23. Bouali H, Kolinsky R. Source evaluation: Components and impacts. Thinking Skills and Creativity [Internet]. 2023 Mar 1 [cited 2025 Jul 16];47:101250. Available from: https://www.sciencedirect.com/science/article/pii/S1871187123000202

24. Johnson MK, Hashtroudi S, Lindsay DS. Source monitoring. Psychological Bulletin. 1993;114(1):3–28.

25. Wiley J, Goldman SR, Graesser AC, Sanchez CA, Ash IK, Hemmerich JA. Source evaluation, comprehension, and learning in Internet science inquiry tasks. American Educational Research Journal. 2009;46(4):1060–106.

26. Polyn SM, Norman KA, Kahana MJ. A context maintenance and retrieval model of organizational processes in free recall. Psychol Rev. 2009 Jan;116(1):129–56.

